# Genome Wide Association Metanalysis Of Skull Bone Mineral Density Identifies Loci Relevant For Osteoporosis And Craniosynostosis

**DOI:** 10.1101/2021.11.01.21265592

**Authors:** Carolina Medina-Gomez, Benjamin H. Mullin, Alessandra Chesi, Vid Prijatelj, John P. Kemp, Chen Shochat-Carvalho, Katerina Trajanoska, Carol Wang, Raimo Joro, Tavia E. Evans, Katharina E. Schraut, Ruifang Li-Gao, Tarunveer S. Ahluwalia, M. Carola Zillikens, Kun Zhu, Dennis O. Mook-Kanamori, Daniel S. Evans, Maria Nethander, Maria J. Knol, Gudmar Thorleifsson, Ivana Prokic, Babette Zemel, Linda Broer, Natasja van Schoor, Sjur Reppe, Mikolaj A. Pawlak, Stuart H. Ralston, Nathalie van der Velde, Mattias Lorentzon, Kari Stefansson, Hieab H.H. Adams, Scott G. Wilson, M. Arfan Ikram, John P. Walsh, Timo A. Lakka, Kaare M. Gautvik, James F Wilson, Eric S. Orwoll, Cornelia M. van Duijn, Klaus Bønnelykke, Andre G. Uitterlinden, Unnur Stykársdóttir, Timothy D. Spector, Jonathan H Tobias, Claes Ohlsson, Janine F. Felix, Hans Bisgaard, Struan F.A. Grant, J. Brent Richards, David M. Evans, Bram van der Eerden, Jeroen van de Peppel, Cheryl Ackert-Bicknell, David Karasik, Erika Kague, Fernando Rivadeneira

## Abstract

Skull bone mineral density (SK-BMD) provides a suitable trait for the discovery of genes important to bone biology in general, and particularly for identifying components unique to intramembranous ossification, which cannot be captured at other skeletal sites. We assessed genetic determinants of SK-BMD in 43,800 individuals, identifying 59 genome-wide significant loci (4 novel), explaining 12.5% of its variance. Pathway and enrichment analyses of the association signals resulted in clustering within gene-sets involved in regulating the development of the skeleton; overexpressed in the musculoskeletal system; and enriched in enhancer and transcribed regions in osteoblasts. From the four novel loci (mapping to *ZIC1, PRKAR1A, ATP6V1C1, GLRX3*), two (*ZIC1* and *PRKAR1A*) have previously been related to craniofacial developmental defects. Functional validation of skull development in zebrafish revealed abnormal cranial bone initiation that culminated in ectopic sutures and reduced BMD in mutated *zic1* and *atp6v1c1* fish and asymmetric bone growth and elevated BMD in mutated *prkar1a* fish. We confirmed a role of *ZIC1* loss-of-function in suture patterning and discovered *ATP6V1C1* gene associated with suture development. In light of the evidence presented suggesting that SK-BMD is genetically related to craniofacial abnormalities, our study opens new avenues to the understanding of the pathophysiology of craniofacial defects and towards the effective pharmacological treatment of bone diseases.

## Introduction

Bone mineral density (BMD) as measured by Dual X-ray Absorptiometry (DXA) is the primary diagnostic marker for osteoporosis and fracture susceptibility in adults. Large scale genome-wide association studies (GWAS) using adult BMD measurements of clinically relevant weight bearing skeletal sites (hip and spine), have been successful in identifying genetic variants that account for a small proportion of the variance in BMD^1; 2^. Also, GWAS of bone characteristics evaluated by other techniques such as peripheral quantitative computed tomography (pQCT) (assessed in the tibia or forearm) ^3^ and ultrasound (assessed in the heel)^4; 5^, have unveiled hundreds of loci influencing bone metabolism. Nevertheless, investigating the origin of BMD variation in the skull has been largely disregarded by genetic studies, possibly viewed as irrelevant to the processes influencing osteoporosis and fracture risk.

Only one previous study in ∼9,500 children investigated the genetic architecture of skull BMD (SK-BMD). The GWAS identified 8 loci, all of which were previously associated with BMD at other skeletal sites^6^, however, genetic correlation analyses showed that SK-BMD has to a large extent a distinct genetic architecture^6^. Another large study of total body BMD within the setting of the GEFOS consortium exists^7^, but SK-BMD was not examined separately. Characterizing the skull as a site-specific phenotype may unravel novel genes and pathways underlying bone biology, specifically those related to mechano-sensing and intramembranous ossification. The cranial bones are under less mechanical strain compared to long bones; it has been estimated that the human fibula is exposed to nearly twice the load that the human skull experiences^8^. As such, calvarial osteocytes are considered to be exceptionally mechanosensitive, as they can maintain their physiological function despite low stimuli from mechanical loads and muscle traction^9^. The study of SK-BMD presents a unique opportunity to unveil aspects of bone mineralization of craniofacial structures. In contrast to the rest of the body, the skull has a dual embryonic origin from cranial neural crest cells (CNCC) and paraxial mesoderm (PM) and ossifies through intramembranous ossification^10^. Thus, the purpose of our study was to identify genetic variants associated with SK-BMD in a large meta-analysis unravelling new genes potentially involved in craniofacial development and disease while also affecting BMD.

## Methods

### SK-BMD GWAS meta-analyses

#### Study Populations

##### Subjects

This study comprised ∼43,800 individuals, taking part in 21 epidemiological studies worldwide (**Table S1-S2**). The majority of these individuals were adults (>18 years, 75%) from cohorts of European background (85%). Written informed consent was provided by all subjects (or their parents in the case of children), and this study was approved by the corresponding Medical Ethics Committee of each participating study.

Skull BMD (g/cm^2^) was measured by DXA following standard manufacturer protocols in each research center (**Table S1**). All individuals included in this study had genome-wide array data imputed to the 1000 genomes phase 1 version 3 (March 2012) reference panel or the combined 1000 genomes and the UK10K reference panels (**Table S1**).

#### Association Analysis

Individual cohorts generated normalized SK-BMD residuals after adjustment for age, weight, height and genomic principal components, either in sex-combined (family-based studies) or in sex-specific (population-based studies) models then combined for analysis. These normalized residuals were tested for association with genome-wide single nucleotide polymorphisms (SNPs) in an additive model.

##### Quality control of SK-BMD association summary statistics

In the first instance, a centralized quality-control procedure implemented in EasyQC^11^ was applied to individual cohort association summary statistics. Cohort-specific errors in phenotype residual transformation or inflation arising from population stratification, cryptic relatedness and genotype biases were evaluated and corrected when necessary. Moreover, variants with missing information, or nonsensical values (e.g., absolute beta estimates or standard errors >10, association P-values >1 or <0; or imputation quality < 0; infinite beta estimates or standard errors); minor allele frequency (MAF) less than 0.5%; imputation quality scores <0.4 (Impute2 info) or <0.3 (Minimac r^2^), were excluded.

##### GWAS meta-analyses

An inverse-variance meta-analysis was carried out in METAL^12^, surveying 19,211,311 markers present in at least three studies after quality control. We applied the conventional genome-wide significance (GWS, P≤5×10^−8^) threshold for SNP discovery.

##### Approximate conditional meta-analyses

Conditional analyses were undertaken based on the meta-analysis results employing an iterative strategy as implemented in GCTA ^13^, using the Rotterdam Study I (n=6,291) dataset as reference for precise calculation of the linkage disequilibrium (LD) between the analyzed markers. We employed this tool to determine: 1) independence of association signals within loci discovered in our study; and 2) independence of the association signals discovered by our meta-analysis from the 570 independent variants, present in our meta-analysis, that have been reported previously in well-powered GWAS of relevant bone traits ^1-4; 6; 7; 14-17^ including the last UKBB estimated-BMD survey ^4^ (**Table S3**).

### Shared Genetic architecture of SK-BMD, fracture and other traits

#### LD score regression analyses

We used LD score regression ^18^ as implemented in the LDhub web interface ^19^ to rule out that our results were a product of residual population stratification or cryptic relatedness and to estimate the SNP heritability of SK-BMD. Likewise, this web utility can estimate the genetic correlation (ρ_g_) between two traits based on GWAS summary statistics^18^. In addition, we assessed the genetic correlation between SK-BMD and other relevant traits not present in the LDhub database, i.e., all type of fracture^20^, handgrip strength ^21^ and different lobar brain volumes^22^.

##### Search for biological and functional knowledge of the identified association regions

Functional mapping and annotation of genetic associations was performed with FUMA ^23^. Also, Combined Annotation Dependent Depletion (CADD) scores for exonic variants were retrieved from this tool as well as annotations to the GWAS catalogue to examine pleiotropic relationships.

#### Enrichment of GWAS variants for regulatory annotations

GWAS Analysis of Regulatory or Functional Information Enrichment with LD correction (GARFIELD)^24^ is a framework of tools used to analyze enrichment for traits and annotations of primarily GWAS results whilst taking multiple confounders into account. We analyzed our variants for enrichment at variable GWS p-value thresholds (i.e., 5 × 10^−5^, 5 × 10^−6^, 5 × 10^−7^, 5 × 10^−8^) against chromatin states and histone modifications of osteoblast signatures (as acquired from multiple sources^25; 26^) and across other cell lines (GM12878, H1HESC, HeLa-S3, HepG2, HUVEC, K562; as provided by GARFIELD). We tested for enrichment of ATAC-Seq (Assay for Transposase-Accessible Chromatin using sequencing) ^5^ and DNase I hypersensitive site (DHS) peaks of osteoblast signatures^27^. In total, we tested for enrichment of 116 annotations spread across all cell lines and features, including ATAC-Seq and DHS marks.

#### DEPICT gene prioritization and pathway annotation

We used DEPICT ^28^ at a GWS threshold, to prioritize genes at the associated regions and highlight important pathways influencing skull mineralization. Enriched gene-sets were grouped based on the degree of gene overlap into ‘meta gene-sets’ using affinity propagation clustering^28^. Visualization was carried out using Cytoscape 3.4.

#### Osteoclast eQTL analysis

In order to identify potential regulatory effects relevant to bone cells for the GWS variants in this study, we performed an expression quantitative trait locus (eQTL) analysis of these variants in a previously generated osteoclast eQTL dataset^29; 30^. A detailed description of the patient recruitment process and laboratory protocols used in this study was recently published^29^. Briefly, gene expression profiles were generated by performing RNA-Seq on osteoclast-like cells differentiated from peripheral blood mononuclear cells (PBMCs) *in-vitro*. These cells were isolated from 158 female patients aged 30 to 70 years for whom genome-wide genotype data imputed to the haplotype reference consortium (HRC) panel release 1.1 was also available. The eQTL analysis was performed, as described in detail previously^29^, on the osteoclast gene expression data normalized using the trimmed mean of M-values method and corrected for total read count by conversion to counts per million using the *edgeR* package in R ^31^, and only included variants with a MAF ≥5%. Models were adjusted for age, RNA-Seq batch and 10 principal components. Each variant was tested for association with the expression of any gene with a transcription start site that fell within a 1 Mb window (*cis-eQTLs*). Correction for multiple testing was performed using the Benjamini-Yekutieli procedure with a false discovery rate (FDR) of 5%. We used two different approaches to analyze this data, namely: 1) Co-localization of GWAS/eQTL association signals ^32^ and 2) Summary-data based Mendelian randomization (SMR) analysis ^33^, as we briefly explain below.

##### Co-localization analysis

Assessment of co-localization, using a Bayesian framework to calculate posterior probabilities to quantify support for five different hypotheses regarding the presence and sharing of causal variants for eQTLs and SK-BMD, was performed using the coloc package in R ^32^.

##### SMR analysis

We attempted to identify genes whose expression levels could potentially mediate the association between SNPs and SK-BMD using the SMR software ^33^. Briefly, this method tests for association between gene expression and a given trait using the most associated eQTL as a genetic instrument. The software performs a SMR test, which uses the top eQTL variant for each gene to identify association signals present in both the GWAS and eQTL datasets. The software also performs a heterogeneity in dependent instruments (HEIDI) test, which tests for heterogeneity in the *cis-eQTL* region to determine if there is a single causal variant underlying the GWAS and transcriptomic signals. A significant HEIDI test at a particular locus indicates the presence of heterogeneity for the two datasets, indicating the association signals are less likely to be driven by the same causal variant. We followed this approach directly in the osteoclast-specific eQTL dataset from 158 participants described above ^29^, and also in the better-powered eQTL study in whole blood reported by Westra *et al*. ^34^. The genotype data from the osteoclast eQTL cohort was used as the reference panel in this analysis for estimation of LD. The analysis only included genes with at least one eQTL association at P≤5×10^−8^, with correction for multiple testing performed using the Bonferroni method.

#### Osteoblast ATAC-seq and Capture-C

A method to characterize the genome-wide interactions of all human promoters in an osteoblast model using ATAC-seq and high-resolution Capture-C was recently developed^35^. In short, a custom Agilent SureSelect RNA library targeting *DpnII* restriction fragments overlapping 36,691 promoters of protein-coding, noncoding, antisense, small nuclear (sn)RNA, micro (mi)RNA, small nucleolar (sno)RNA and long intergenic noncoding (linc)RNA genes was designed. Then, genome-wide, promoter-focused high-resolution Capture-C was applied to primary human mesenchymal progenitor cell (MSC)-derived osteoblasts. Also, ATAC-seq open chromatin maps from the same samples were generated to determine informative proxy SNPs for each of the SK-BMD loci. The intersection of these two datasets provides an indication of the genes being targeted by the SNPs associated with SK-BMD and thus, more likely to be mediating the association signal. Significant interactions were called using the CHiCAGO pipeline^36^.

### Gene expression in human bone and knockout animal models

#### Human bone tissue - OSTEOGENE Project

Eighty-four biopsies from female iliac bone donors and sixty-six subchondral bone fragments from women undergoing hip replacement surgery due to hip fracture or osteoarthritis were subjected to transcriptomic analysis. Detailed description on sampling and characteristics of these women can be found elsewhere^37^. For the bone fragment collection, standardized extraction from a 1 cm^2^ area of the caput was performed during surgery and frozen in liquid nitrogen. The frozen bone fragments were then pulverized in a mortar followed by RNA extraction with a Trizol reagent (Life Technologies, Gaithersburg, MD) and further purification using a RNeasy kit (Qiagen). RNA from all the bone samples (biopsies and surgical fragments) were then sequenced in a single batch using TruSeq RNA Library prep kit V2 (Illumina) and single indexed adapters. Paired-end sequencing with 2 × 50 bp was performed using the Illumina Hiseq2000 platform to obtain at least 6,000,000 reads per library. Transcript level expression values were then created using an in-house pipeline utilizing Picard tools (http://broadinstitute.github.io/picard/), GATK^38^ and featureCounts^39^. Sample-donor annotation concordance was ensured. No libraries had fewer than 100,000 reads. Expression data was then quantile normalized and genes not expressed in at least 75% of libraries were excluded.

#### Human mesenchymal stem cells

Human bone marrow derived mesenchymal stem cells [(hMSC), Lonza Group Ltd., Basel, Switzerland] were seeded in 12-well plates (5×10^3^ cells per cm^2^) and differentiated into osteoblasts (using α-Mem pH7.5, 10% heat inactivated fetal calf serum (FCS), 100 nM Dexamethasone and 10 mM β-glycerophosphate). As mentioned in the datasheet provided by the company, cells were authenticated by FACS analyses for the presence of surface markers CD105, CD166, CD29 and CD44 and the absence of CD14, CD34 and CD45. In addition, osteogenic, adipogenic and chondrogenic differentiation was demonstrated by alizarin red S staining, oil-red-O staining and collagen II staining, respectively. The human MSCs were tested negative for mycoplasma, both by the company and in-house during the culture experiments described in this manuscript. Total RNA was isolated using Trizol (Life Technologies, Carlsbad, CA, USA) as described previously^40^. This data can be accessed through the NCBI-GEO (Accession number GSE80614).

#### Animal model surveys

Genes prioritized either by location, function (DEPICT), eQTL or Capture C analyses were searched in both the Mouse Genome Database^41^ (MGD; http://www.informatics.jax.org) and the International Mouse Phenotyping Consortium^42^ (IMPC, https://www.mousephenotype.org/) surveys.

#### Gene expression in murine bone cells

Gene expression profiles of candidate genes were examined in primary mouse osteoblasts undergoing differentiation and bone marrow derived osteoclasts. To study murine osteoblasts, pre-osteoblast-like cells were obtained from neonatal calvaria collected from C57BL/6J. Next Generation RNA sequencing using an Illumina HiSeq 2000 was used to evaluate the transcriptome every two days from day 2 to 18 days post osteoblast differentiation^7^. Expression of genes in murine osteoclasts was determined using publicly available data obtained using Next-Gen RNA-sequencing applied to bone marrow derived osteoclasts obtained from 6-8-week-old C57BL/6 mice^43^. All procedures and use of mice for the neonatal osteoblast expression studies were approved by the Jackson Laboratory Animal Care and Use Committee (ACUC), in accordance with NIH guidelines for the care and use of laboratory animals.

#### Assessment of gene function in skull bone of mutant Zebrafish

##### Zebrafish CRISPR/Cas9 injections

We used three synthetic guide (g) RNAs (ordered as crispr (cr) RNAs, Sigma) targeting the most plausible orthologs of all three genes (*zic1; atp6v1c1a and atp6v1c1b; prkar1aa and prkar1ab*) (**Table S4**) of the four novel loci (**Figure 1**). The signal mapping to chromosome 10, was not followed up given the large distance to the closest gene and the complex recombination pattern of the region (**Figure 1**). For *zic1*, we used three crRNAs (2pg), while for the other genes with more than one gene in zebrafish, we concomitantly targeted all orthologs using six crRNAs. crRNAs were incubated with trans-activating (tra) crRNA (10pg) and GeneArt Platinum Cas9 nuclease (Invitrogen) prior to injections. Injections (1nl) were performed into 1-cell stage of embryos of the osteoblast reporter lines Tg(*osx:NTR-mCherry*)^44^ or Tg(*Ola*.*Sp7*:NLS-*gfp*)^45^. Osterix (Osx or Sp7) is a marker of osteoblast maturation. To validate CRISPR efficiency (90%), DNA was extracted from 12 individual injected larvae at 5dpf (days post-fertilization), followed by PCR amplification using FAM-M13F primer and gene-specific primers, with each forward primer containing an M13 tail (**Table S5**). PCRs were submitted to fragment length analysis (ABI 3500)^46^. Controls were injected with Cas9 protein and SygRNA^®^ SpCas9 tracrRNA (10pg) (Merck).

**Figure 1.**
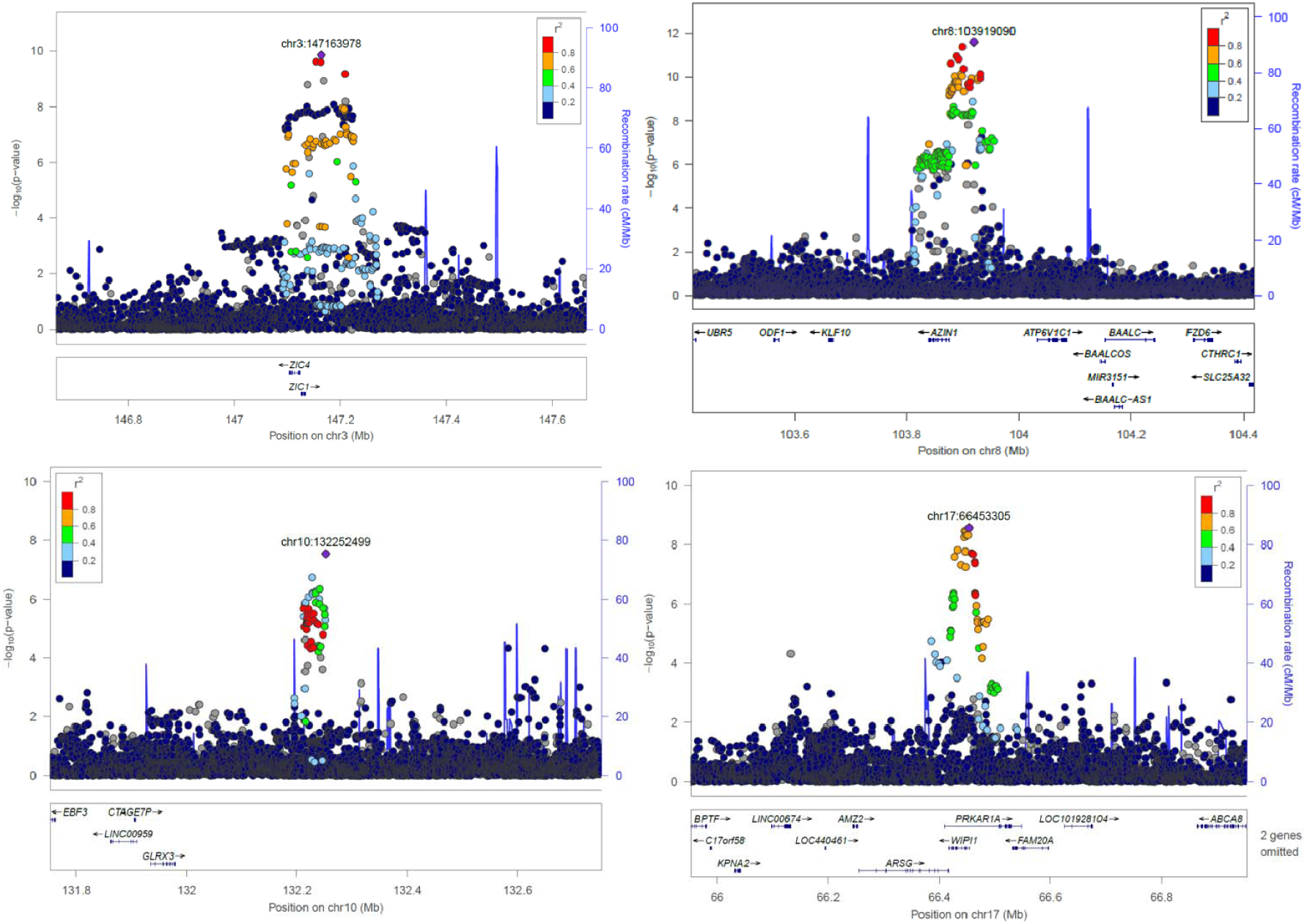
Regional plots for the four novel loci associated with SK-BMD (P<5×10). Circles show GWAS meta-analysis P-values and position of SNPs for the overall meta-analysis (N=43,800). Different colors indicate varying degrees of pair-wise linkage disequilibrium with the top marker (1000 Genomes – CEU population

##### Bone assessments in Zebrafish

Juvenile Tg(*osx:NTR-mCherry*) or Tg(*Ola*.*Sp7*:NLS-*gfp*) crispants (G0s, injected fish) were briefly anesthetized using tricaine (MS222) followed by live imaging of their skulls using a Leica Fluorescent microscope and LasX software. *Ex vivo* Alizarin Red staining was performed in two-month fish. Animals were euthanized and fixed in 4% PFA followed by an acid free bone staining, as previously described^47^. Animal experiments were ethically approved by the University of Bristol Animal Welfare and Ethical Review Body (AWERB) and conducted under UK Home Office project license.

##### Zebrafish Micro Computed Tomography (μCT)

A total of 21 two-months old fish were fixed in 4% PFA for 7 days, dehydrated in 70% ethanol solution and scanned using a 1172 SkyScan *μ*CT scanner (Bruker, Kontich, Belgium) at pixel size of 12 *μ*m (scan settings 49 kV, 100 *μ*A, filter Al 0.25mm). Images were reconstructed using NRecon Software (Bruker). BMD was measured from whole-body and skull-only regions using CTan Software (Bruker), previously calibrated to the phantoms with known mineral density (0.25 and 0.75⍰g.cm^−3^ hydroxyapatite, Bruker). BMD was compared between groups displaying similar body length. *prkar1a* crispants were smaller and therefore were compared with controls matched by size (standard length). A representative skull from each studied group was re-scanned at a pixel size of 3 *μ*m. Amira 6.0 (FEI) was used to generate 3D volume renders with the same parameters for each group.

## Results

### SK-BMD is a highly polygenic trait and unveils 4 novel BMD loci

Our meta-analysis of SK-BMD GWAS summary statistics (N=∼43,800) identified 79 independent signals mapping to 59 loci (**Figures S1-S2, Table S6**). The variants associated with SK-BMD at genome-wide significant (P≤5×10^−8^) level explain 12.5% of the SK-BMD variance. The overall genomic inflation factor (*λ*) of 1.09, (**Figure S3**), and of common variants (0.2>MAF<0.5, *λ*=1.17) is compatible with trait polygenicity (LD score regression intercept = 1.022). Besides establishing independent SNPs associated with SK-BMD, the conditional analysis was used to establish novelty of the associations identified by our analysis (Table S7, Figure S4). Signals mapping to 4 different loci i.e., 3q24 [*ZIC1*/ZIC4], 8q22.3 [*AZIN1/ATP6V1C1*], 10q26.3 [*GLRX3*], and 17q24.2 [*PRKAR1A*/*WIPI1*] have not been reported previously in GWAS meta-analyses of bone traits (**Table 1, Figure 1**). Moreover, association signals mapping to 1p13.2 [*ST7L*]; 1q23.3 [*LOC100422212*]; 5q13.2 [*FOXD1*], 6p12.1 [*LRRC1*], 6p25.1 [*RREB1*], 6q13 [*CD109*], 6q23.1 [*L3MBTL3*], 8q23.1 [*EMC2*], 12p11.22 [*CCDC91*], 12q12.12 [*ADCY6*], 12q22 [*SOCS2*], 17p13.3 *[NXN], and* 17q24.3 *[MAP2K6, KCNJ16, KCNJ2]* are for the first time reported as associated with DXA-derived BMD (**Figure S1**). Novel independent signals in previously identified BMD loci were also detected at 1p31.3 [*WLS*], 6q22.33 [*CENPW*] and 13q14.11 [*TNFSF11*], suggesting the existence of distinct regulatory elements shaping skull-BMD variation. Further, 25 non-synonymous exonic and one stop-gain variants were observed across 16 different genes (**Table S8**). Of these, nine are classified as belonging to the 1% most deleterious variants in the human genome (CADD score > 20), and map to genes with well-known roles in bone biology, including: *LGR4*^*48*^, *LRP5*^*49*^, *WNT16*^*50*^, *CPED1*^*15*^, *LRP4*^*51*^, *BDNF*^*52*^, *PRKAG1*^*53*^, *ADCY6*^*54*^.

**Table 1.**
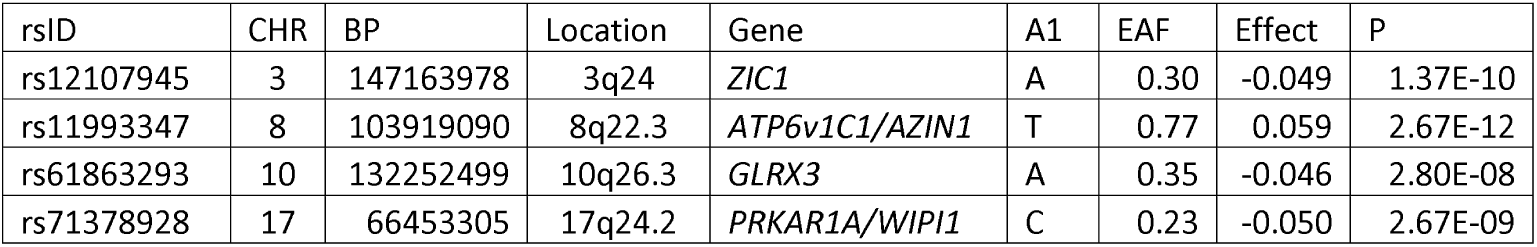
Index SNPs of novel loci associated with BMD. Variants associated with SK-BMD in the meta-analysis that map outside +/- 500 Kb of known index SNPs of genetic associations with different bone traits. Genomic coordinates are on build 37 of the human genome. Notes refer to annotation based on the closest gene. Effect sizes and allele frequencies (EAF) are reported for the A1 allele

### SK-BMD is correlated to fracture and other traits

SNP-heritability of SK-BMD was estimated to be 0.306 (SE 0.028). Positive genetic correlation of SK-BMD with other site-specific BMD ranged between 0.347 and 0.765, while it was lower and negative with fracture risk (ρ_g_ =-0.38; SE 0.06) (**Table S9**). Besides a positive correlation with sitting height (ρ_g_= 0.275; SE 0.058), no other significant genetic correlations after multiple-testing correction (53 traits, P≤9.4×10^−4^) were identified.

### Biological and functional knowledge of the genes in SK-BMD associated loci

#### SK-BMD variants are enriched in osteoblast functional signatures

In total, 116 functional signatures were tested for enrichment using GARFIELD (Bonferroni corrected P≤ 4.31 × 10^−4^). Variants associated at GWS threshold showed clear enrichment for enhancer, weak enhancer and transcribed regions in osteoblasts (**Figure 2**). Concordantly, trimethylation of the 27^th^ lysine residue of the 3^rd^ histone (H3K27me3) and trimethylation of the 9^th^ lysine residue of the 3^rd^ histone (H3K9me3) were not enriched, a result in line with lack of enrichment for repressed chromatin states. Similarly, variants at GWS threshold showed a uniform pattern of lack of enrichment in non-osteoblast cell lines. In contrast, variants were enriched for ATAC-Seq signatures in osteoblasts across all genome-wide p-value thresholds (**Table S10**).

**Figure 2.**
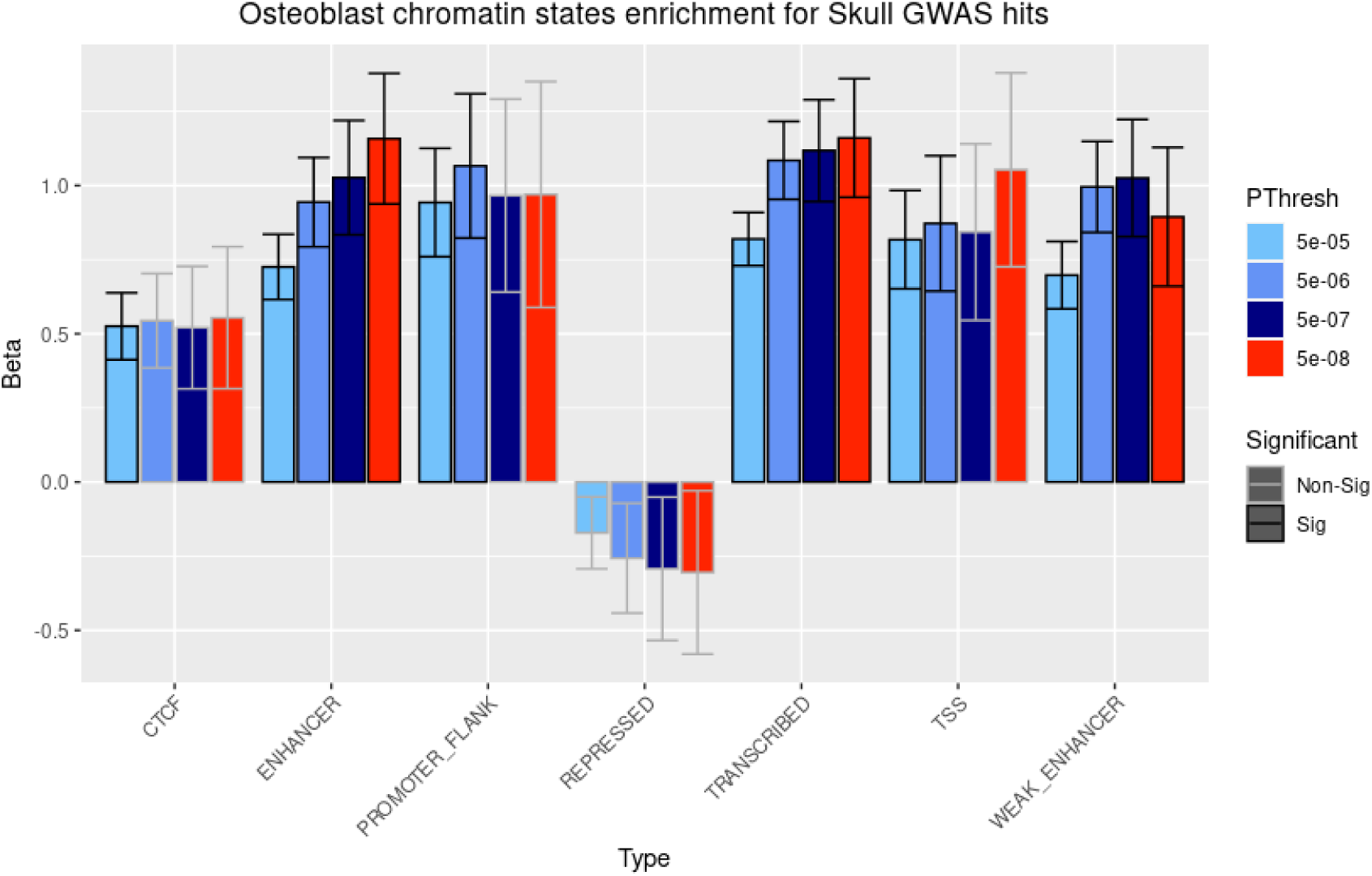
GARFIELD results for chromatin states enrichment analysis in osteoblasts. Enrichment significance was defined at P< 4.31×10^−4^. Bar-plot fill colors represent p-value GWS threshold of variants included in the analysis. Bar-plot outline colors represent significance of the enrichment. GWS variants were enriched for enhancer, weak enhancer and transcribed osteoblast regions.

#### SK-BMD variants hint to genes relevant in craniofacial development

Based on the GWAS meta-analysis, 22 genes were prioritized (FDR<0.05) (**Table S11**). Cells and tissues from the musculoskeletal system presented the largest enrichment of gene expression within the associated loci (**Figure 3**). These genes were overrepresented in 194 pathways clustered in 28 ‘meta gene-sets’ (**Table S12**). These clusters are almost exclusively involved in development of skeletal and other cranial components (i.e., bony ear, jaw) (**Figure 3**), suggesting that our analyses are capturing genes involved in craniofacial development.

**Figure 3.**
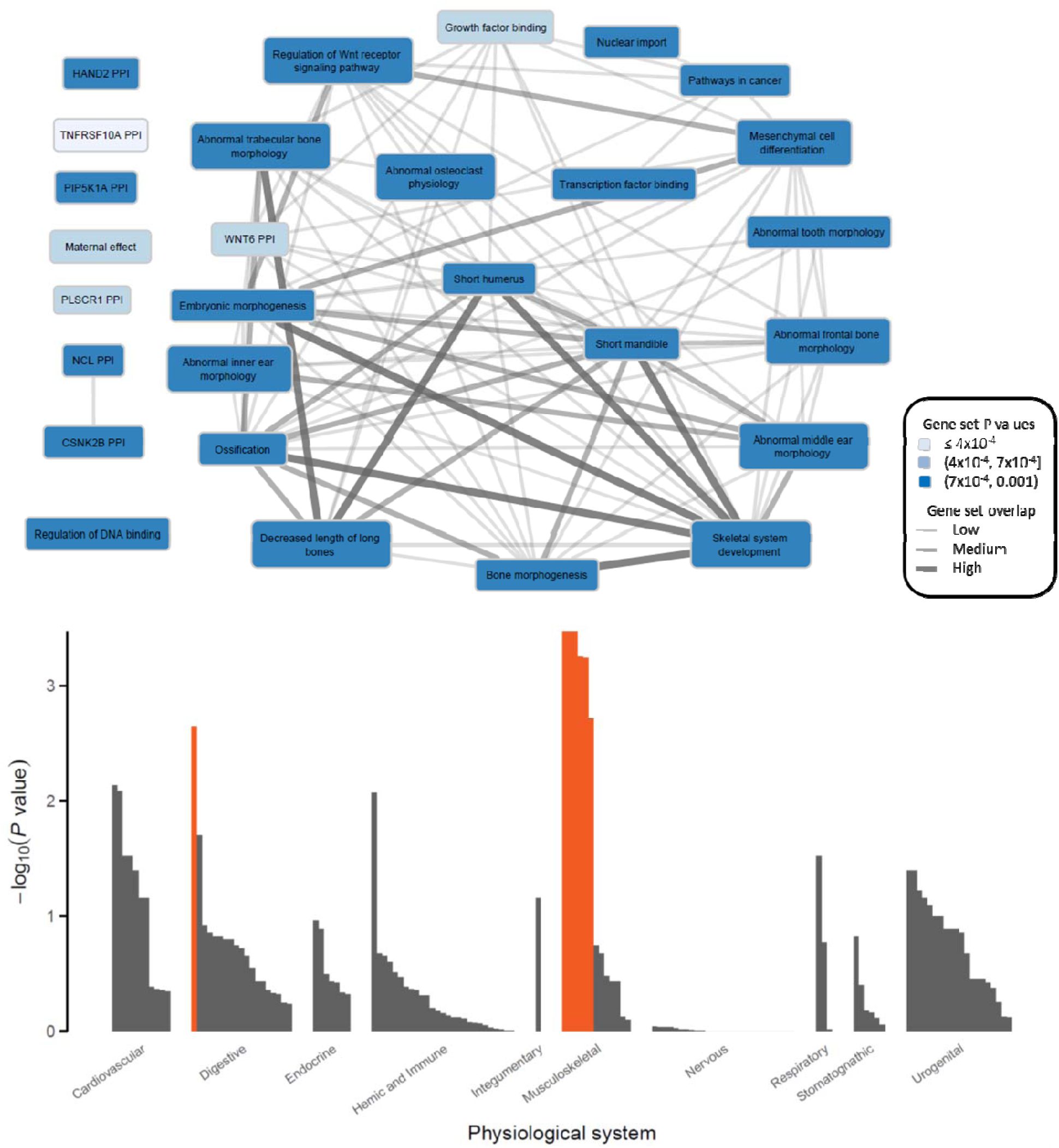
DEPICT results for gene-set and cell/tissue enrichment analyses. **Top panel:** Meta gene-sets were defined from similarity clustering of significantly enriched gene sets (FDR<5%). Each Meta gene-set was named after one of its member gene sets. The color of the Meta gene-sets represents the P value of the member set. Interconnection line width represents the Pearson correlation ρ between the gene membership scores for each Meta gene-set (ρ < 0.3, no line; 0.3 ≤ ρ < 0.45, narrow width; 0.45 ≤ ρ < 0.65, medium width; ρ ≥ 0.65, thick width). **Bottom panel:** Bars represent the level of evidence for genes in the associated loci to be expressed in any of the 209 Medical Subject Heading (MeSH) tissue and cell type annotations. Highlighted in orange are these cell/tissu types significantly (FDR<5%) enriched for the expression of the genes in the associated loci

#### SK-BMD variants are not associated with osteoclast gene expression

##### Co-localization analysis

Osteoclast eQTL association signals were surveyed based on the 79 independent SK-BMD variants. There were 6 eQTL associations that remained significant after correction for multiple testing (FDR ≤5%), mapping to *ST7L* (1q13.2), *REEP5* (5q22.2); *ING3* (7q31.31), *RIC8A* (11p15.5), and *CDC42* and *LINC00339* (1p36.12). Moreover, when assessing co-localisation of GWAS/eQTL association signals for these six signals, in two instances (*RIC8A* and *REEP5*) the results suggested the presence of a single causal variant driving both the GWAS and eQTLs associations. (**Table S13**).

##### SMR analysis

Analysis of the SK-BMD GWAS dataset with the blood eQTL database ^34^ resulted in 15 genes with significant associations in the SMR test (P_SMR_≤0.05/5912=8.5×10^−6^; 5,912 genes included in the analysis) (**Table S14**). For 7 of these, there was no evidence of heterogeneity in the association signals from the two datasets (P_HEIDI_≥0.05), suggesting that the GWAS and eQTL association signals may be driven by the same causal variant rather than by different variants mapping to the same locus: 1p13.2 (*MOV10*); 6p21.1 (*SUPT3H*); 8q22.3 (*AZIN1-AS1*); 12q23.3 (*TMEM263*); 14q24.3 (*EIF2B2* and *MLH3*); 20q12 (*MAFB*). SMR analysis of the osteoclast eQTL (1,077 genes included in the analysis)^29^ and SK-BMD datasets resulted in significant (P_SMR_≤0.05/1077=4.6×10^−5^) SMR statistics for *LINC00339* (1p36.12) and *TMEM8A* (16p13.3), neither of which were present in the blood eQTL dataset. However, the results from the HEIDI analysis did suggest the presence of heterogeneity (different SNPs in LD responsible for the GWAS and eQTL signals) at these loci (P_HEIDI_<0.05). Interestingly, *EIF2B2* showed a suggestive SMR statistic in the analysis of the osteoclast eQTL and SK-BMD datasets (P =7.0×10^−5^), which was directionally consistent with the results obtained in blood. However, there is evidence of heterogeneity also at this locus. Of note, this locus was only suggestive in the GWAS meta-analysis.

#### SK-BMD variants are implicated in the regulation of genes in osteoblasts

In order to further map putative causal SNPs at the SK-BMD GWAS loci to their target effector genes, we employed a high-resolution genome-scale, promoter-focused Capture-C based approach coupled with ATAC-seq in human primary mesenchymal stem cell (MSC) -derived osteoblasts ^55^. Capture-C detects interactions in 3D space between distant regions of the genome, while ATAC-seq provides a genome-wide map of open chromatin in the same cell type. After filtering out interactions between proxy SNPs (r^2^>0.4) and promoters not in open chromatin, our results implicate the following genes: *C1orf105 [1q24*.*3]; TNFRSF11B/COLEC10 [8q24*.*12]; CTTNBP2NL, MIR4256* and WNT2B [1p13.2]; *DMP1* and *SPARCL1* [4q22.1]; *LOC101927839* and *SMG6* [17p13.3]; *ODF1* [8q22.3]; *RUNX2* [6p21.1]; *SMAD9* [13q13.3]; *TCF7L1* [2p11.2]; *FOXD1-AS1* [5q13.2]; and *WNT4* [1p36.12], and some long non coding RNAs (see **Table S15**).

#### SK-BMD implicated genes are relevant to bone biology

We here summarize the lookups for 56 protein coding genes, that were prioritized based on localization and DEPICT results and which had mouse orthologs. Although expression of these genes in human bone related cells observed no enrichment, most of these genes (91%) were expressed in murine calvaria osteoblasts. Only five genes were not: *C1orf105, Odf1, Mepe, Supt3h* and *Wnt1*. However, expression of the latter two genes was observed in murine osteoclasts. Moreover, 75% (42/56) of these genes have been subjected to knockdown in mouse models and about half showed a bone relevant phenotype reported either in MGI or IMPC (**Table S16**). Whereas, in the human bone biopsies 93% (52/56) of the genes were expressed with the exception of four (i.e, *ODF1, COLEC10, MLL2, TMEM263*). For the four novel loci, the closest genes to each of the GWAS signals were found to be expressed in murine osteoblasts and also in human bone tissue (biopsies and fragments). We did not find detectable expression of *ZIC1* in an in-depth gene expression analyses of osteoblast differentiating human bone-marrow derived mesenchymal stromal cells (hMSCs) and osteoclast differentiating peripheral blood mononuclear cell (PBMCs) (**Figure S5**). Conversely, *PRKAR1A* was highly expressed in all experiments. Overall, expression of this gene tends to reduce within the first day of osteoblast differentiation and then normalizes back to basal levels, whereas in osteoclasts the initial high levels were reduced during proliferation, differentiation, and fusion of the cells, and normalize around day 14 when the osteoclasts mature (**FigureS6**). Expression of *ATP6V1C1* is upregulated at the onset of the extracellular matrix mineralization and at the start of the PBMC to osteoclast differentiation (**Figure S7**).

##### Zebrafish studies identifies a role of the *ATP6V1C1* in suture patterning

Three out of the four novel loci (*ZIC1*/ZIC4, *AZIN1/ATP6V1C1, GLRX3*, and *PRKAR1A*/*WIPI1*) harbored genes previously involved in bone biology: craniosynostosis (*ZIC1*)^56^, osteoclast proton pump (*ATP6V1C1*) ^57^, and intramembranous ossification (*PRKAR1A*)^58^. We questioned whether SK-BMD candidate genes from novel loci would have any functional role in craniofacial development. Zebrafish have emerged as advantageous animal models for skeletal diseases, including osteoporosis and craniosynostosis^59; 60^. Zebrafish are particularly suitable for craniofacial development, because they permit non-invasive *in vivo* analysis of skull and sutures during their formation^61^. Moreover, the CRISPR/Cas9 technology allows the generation of biallelic mutations in zebrafish G0s (crispants), phenotypically recapitulating skeletal phenotypes of knockouts and allowing to rapidly assess the function of candidate genes^62; 63^. We used CRISPR/Cas9 to induce loss-of-function mutations in *zic1*, and concomitantly in both zebrafish orthologs of *ATP6V1C1* (*atp6v1c1a* and *atp6v1c1b*) and of *PRKAR1A* (*prkar1a*a and *prkar1a*b) (**Figure 4B**). We monitored the development of the sutures of each of the studied groups (control n= 200, *zic1* n= 64, *atp6v1c1 (a+b)* n = 34, *prkar1a (a+b)* n= 35) using osteoblast reporter lines Tg(*Ola*.*Sp7*:*NLS-GFP*) or Tg(*osx:NTR-mCherry*) (**Figure 4A and B**). *Zic1* zebrafish crispants showed abnormal skull development and bone growth (14/64, 22%) with open calvaria (caput membranaceum) or central foramina that culminated in suture mis-patterning, validating our experimental design (**Figure 4C**). Similarly to *zic1* crispants, *atp6v1c1(a+b)* crispants displayed abnormal skull development (11/34: 32%), with abnormal bone condensation, parietal foramina, uneven bone growth (frontal and parietal) and mis-patterning of the sutures. *Prkar1a(a+b)* crispants showed slower and asymmetric bone growth, detected by small cavities in the skull during development (10/35), while ectopic sutures were not observed (**Figure 4C**). Crispants were compared to 200 controls, that displayed normal bone growth and suture pattern. *Ex vivo* evaluation of the skull using Alizarin Red staining showed identical results (**Figure 4C**). Next, we performed micro-computed tomography to analyze BMD (**Figure 4D**). Comparing fish of similar standard lengths, we detected significant reduction in BMD of *zic1* (p= 0.0025 and p=0.007, for skulls and whole-bodies, respectively) and *atp6v1c1(a+b)* (p= 0.066 and p= 0.015, respectively) crispants, while increased BMD in *prkar1a*(a+b) crispants (p= 0.023 and p=0.241, for skulls and whole-bodies, respectively) (**Figure 4E-H**). Remarkably, BMD variation in the skull and whole-body followed the same trend in all groups, suggesting that genes affecting skull development, in some degree, also regulate BMD of the axial skeleton.

**Figure 4.**
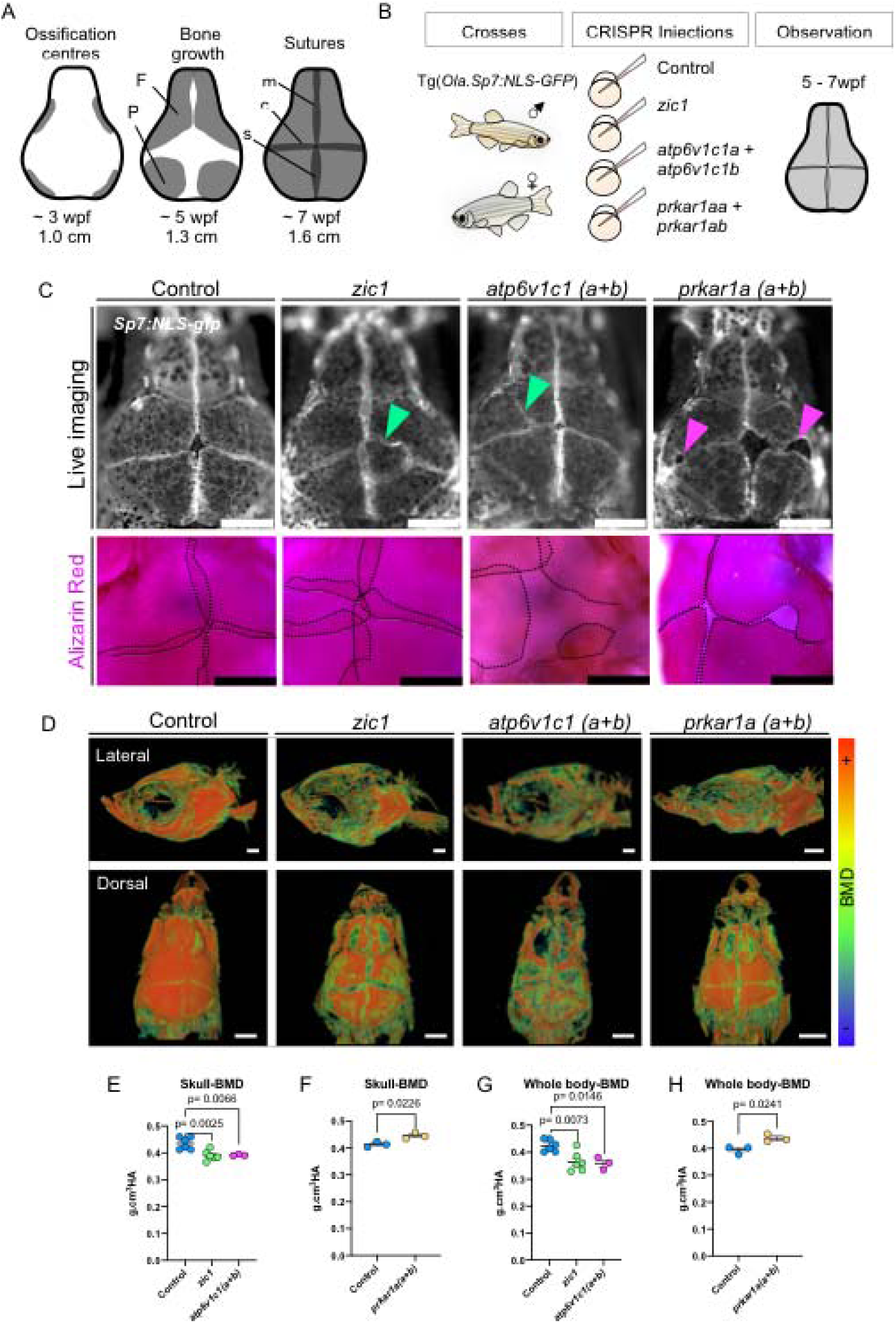
Rapid functional evaluation of novel BMD associated genes in zebrafish identifies a role of novel gene *atp6v1c1* in skull development and a role of *prkar1a* in bone growth. A) Schematic of skull formation in zebrafish. Four ossification centres (grey) are formed by condensation of osteoblasts in the periphery of the skull at around 3 weeks post-fertilization (wpf). Ossification planes grow towards the centre of the skull forming the frontal (F) and parietal (P) bones, completely covering the brain at 7wpf and forming the metopic (m), coronal (c) and sagittal (s) sutures. B) Schematics of the zebrafish crispant experiments. Fish carrying osteoblast reporter line Tg(*Ola*.*Sp7*:NLS-*gfp*) were crossed, and embryos at one cell stage were injected with the CRISPR/Cas9 system targeting respective genes. Observations were carried out *in vivo* during skull/suture formation (5-7wpf). Scale bars = 500um. C) Live imaging of skulls and ex vivo Alizarin Red Staining of control, and *zic1, atp6v1c1 (a+b)* and *prkar1a* (a+b) crispants. Ectopic sutures are indicated with green arrowheads in *zic1* and *atp6v1c1 (a+b)* crispants. Sutures were outlined with a dashed line in Alizarin Red pictures. Cavities in the skull are indicated with magenta arrowheads in *prkar1a* (a+b) crispant. Scale bars = 500um. D) 3D renders micro-computed topographies (*μ*CTs) images of the skull. Images were colour-coded to show BMD variation. E and F) BMD measurement from skulls only from fish of similar standard size (1.8cm). G and H) BMD measurement from whole skeleton from fish of similar standard size (1.6cm). One-way ordinary ANOVA, multiple comparisons test (E and G). Nonparametric, Two-tailed, T-tests (F and H). Data are mean±SD. All P-values are indicated. Graphs were generated in Prism 8.

## Discussion

This meta-analysis of skull BMD comprising up to 43,800 individuals emerges as the first large-scale GWAS in non-pediatric populations focusing specifically on the BMD variation of the skull. Skull BMD constitutes a trait relevant for the study of osteoporosis and fracture risk, as shown by the identification of GWAS signals previously discovered in up to ten times larger GWAS, mapping to genes displaying enrichment for expression in musculoskeletal-related cells and tissues, as well as in enhancer and transcribed regions in osteoblasts. Three genes from the novel loci were screened in zebrafish: *zic1, atp6v1c1* and *prkar1a. Zic1* and *atp6v1c1* crispants displayed low skull BMD and abnormal suture patterning, whereas *PRKAR1A* was involved in calvaria growth and *prkar1a* crispants showed high BMD. We demonstrated that *ATP6V1C1* is not only implicated in bone resorption processes but also in suture patterning and could potentially play a role in craniosynostosis.

Common genetic variants have been shown to explain a greater proportion of variance in BMD at the skull than at appendicular skeletal sites in children^6^. Here, we report a moderately high genetic correlation between SK-BMD and other skeletal sites, attributable to the genetic component controlling systematic processes of bone development. However, we also describe other genetic factors (as denoted by the novel signals in this study) that seem to have a predominant role in the mineralization of skull bones and as such were not detected in studies with a sample size almost 10 times larger^5^.

Two of the leading SNPs from the four novel loci identified by this GWAS, mapped to genes involved in intramembranous ossification (*PRKAR1A*), and neural crest development^64^ and suture patterning^58^ (*ZIC1*). *ZIC1* has also been shown to play an important role in shear flow mechanotransduction in osteocytes^65^ and described to harbor mutations resulting in craniosynostosis^56^. The remaining two GWS signals are intergenic. One of them maps to 8q22.3 in close vicinity to *ATP6V1C1*, an essential component of the osteoclast proton pump^57^. The other signal maps to 10q26.3, located ∼2.7 Mb upstream of the closest protein coding gene (*GLRX3*) involved in iron homeostasis^66^.

While GWS SNPs showed enrichment in enhancers and transcribed regions only in osteoblasts and not in any of the other cells interrogated, the identification of the underlying causal gene at the different associated loci was elusive. Gene prioritization was attempted using different lines of evidence coming from *in-silico* datasets (i.e., DEPICT, CADD scores), chromatin conformation in osteoblasts, eQTLs in osteoclasts, expression in murine and human bone cell lines, and literature evidence. In the four novel loci and in general, the overlap of these lines of evidence was not overwhelming.

*Zic1* was found to be expressed in murine calvarial osteoblasts, however, evidence of expression in the interrogated human cells was not found. This could be explained by the low expression of transcription factors in the cell which is not well-captured by microarray data^67^. Indeed, expression of *ZIC1* was detected in human hip-bone samples using RNA sequencing, but at very low levels.

Gain-of-function mutations in *ZIC1* cause craniosynostosis (premature fusion of the cranial sutures), resulting in restriction of normal growth of the skull, face and brain^58; 68^. Whilst, loss-of-function mutations in *ZIC1* have been associated to calvarial foramina and lack of skull bones (caput membranaceum)^69^, phenotype mirrored by our experiments. The importance of ZIC1 in suture patterning is corroborated by the phenotypic abnormalities of our zebrafish *zic1* crispants. Our data demonstrates that *zic1* is involved in regulation of skull bone condensation, growth and mineralization, providing functional validation to one of the discovered GWAS loci and exquisite comparative base to test other associated candidate genes. Notably, a recent GWAS of parietal brain volume identified a SNP in the *ZIC1* locus^22^. However, the top associated SNP (rs2279829) located 57kb from our SK-BMD leading SNP (rs12107945) is in relatively-low linkage disequilibrium (r^2^=0.14). SNPs in other known BMD loci, such as *CENPW* or *IDUA*, showed also association signals with lobar brain volumes. Although dynamic changes in brain and skull shape track one another during development showing their correspondence, our genetic correlation analysis did not show evidence of significant shared genetic control between brain volumes and skull mineralization.

We additionally tested *atp6v1c1* and *prkar1a* in zebrafish. *Atp6v1c1* crispants revealed a similar phenotype of that observed for *zic1* mutants, including abnormal osteoblast condensation at the ossification centres, abnormal bone growth and ectopic sutures. Recent findings demonstrate that craniosynostosis genes (i.e., *FGFR3*^70^ and *TWIST1* and *TCF12*^*71*^) when mutated in zebrafish might lead to abnormal calvaria bone growth and suture mis-patterning, reinforcing a potential role of *ATP6V1C1* in craniosynostosis. *ATP6V1C1* is a subunit of the V-ATPase complex, which regulates pH by pumping cytosolic protons into intracellular organelles^72^ and has an essential function in osteoclast mediated bone resorption^57^. Mutations in *ATP6V0A3* (*TCIRG1*), another member of V-ATPAses, lead to osteopetrosis^73^. Remarkably, our results also indicate a role of *ATP6V1C1* during osteoblast differentiation, more specifically at the onset of extracellular matrix mineralization, therefore, pointing to a novel direct or indirect function of the gene. On the other hand, loss-of-function mutations in *prkar1a* although showing an increased BMD phenotype, did not display any abnormality on suture patterning.

In craniosynostosis, exacerbation of osteoblast differentiation at the osteogenic fronts of the cranial plates can lead to abnormal extracellular matrix secretion and bone deposition resulting in premature fusion of the sutures^74^. This suggests that genes involved in the regulation of osteoblast differentiation may play an important role in suture patency and fusion. Therefore, it is not surprising that numerous genes identified in BMD associated loci overlap with those identified as causative of craniosynostosis, i.e., *EN1, RUNX2, SOX6, BMP2, JAG1, LRP5, IDUA*^*68*; 75-77^. For instance, haploinsufficiency of *RUNX2* causes cleidocranial dysplasia, a condition which displays opened cranial sutures and lack of mineralization (patent fontanels) in the calvaria^78^. Remarkably craniosynostosis is often caused by gain-of-function mutations, while the opposite (loss-of-function) of the same genes can lead to open foramina, as is the case for *MSX2, RUNX2, ZIC1* and others. Our findings suggest a link between BMD variation and craniosynostosis, postulating genes in BMD loci as potentially implicated in the pathogenesis of craniosynostosis, warranting scrutiny in patients in which the definite mutation underlying the condition has not yet been found.

Altogether, this study is the largest genome-wide survey for association with skull BMD. We demonstrated that the skull provides reliable information on genetic factors that are relevant for the study of osteoporosis and fracture risk. In contrast to other skeletal sites, the study of skull BMD captures distincting elements of intramembraneous ossification, as evidenced by the four novel loci identified. Also, we unveiled a link between BMD, craniosynostosis and skull development, illustrated by the consequences of the loss-of-function *zic1* zebrafish experiments and more importantly, the characterization of *ATP6V1C1* as involved in the regulation of BMD and skull/suture development. The shared genetic architecture between skull BMD and craniosynostosis offers further understanding of the underlying pathophysiology and new avenues for the diagnosis and potential pharmacological treatment of skeletal diseases. Our findings demonstrate the power of our SK-BMD analyses for identification of potential genes involved in craniofacial diseases.

## Data Availability

All data produced in the present study are available upon reasonable request to the authors and will shortly be posted on the GEFOS website: www.gefos.org

## Acknowledgements

The authors would like to thank the many colleagues who contributed to collection and phenotypic characterization of the clinical samples, as well as genotyping and analysis of the GWAS data. Part of this work was conducted using the UK Biobank resource.

Full list of acknowledgements, funding organizations and grants are listed per cohort in the Supplemental Material.

