## Supplemental information for "Genome Wide Association Metanalysis Of Skull Bone Mineral Density Identifies Loci Relevant For Osteoporosis And Craniosynostosis"

### Supplemental data

### COHORTS SHORT DESCRIPTION

Avon Longitudinal Study of Parents and their Children (ALSPAC):

The Avon Longitudinal Study of Parents and their Children (ALSPAC) is a longitudinal population-based birth cohort that recruited pregnant women residing in Avon, UK, with an expected delivery date between 1st April 1991 and 31st December 1992. The initial number of pregnancies enrolled was 14,541. When the oldest children were approximately 7 years of age, an attempt was made to bolster the initial sample with eligible cases who had failed to join the study originally. The total sample size for analyses after the age of seven was 15,454 pregnancies, resulting in 15, 589 foetuses. Of these 14,901 were alive at one year of age. The following manuscript contains the results of ALSPAC individuals who were measured on DXA at the age 9 FOCUS group and have GWAS. Ethical approval for the study was obtained from the ALSPAC Ethics and Law Committee and the Local Research Ethics Committees. Consent for biological samples has been collected in accordance with the Human Tissue Act (2004). Informed consent for use of data collected via questionnaires and clinics was obtained from participants following the recommendations of the ALSPAC Ethics and Law Committee at the time. This cohort is described in detail on the website (http://www.alspac.bris.ac.uk) and elsewhere ^1-3^ and the total body DXA measures and cohort analyzed in the present paper are described in Kemp et al. (2014) ^4^. Please note that the study website contains details of all the data that is available through a fully searchable data dictionary and variable search tool (http://www.bris.ac.uk/alspac/researchers/our-data

Bone Mineral Density in Childhood Study (BMDCS):

The Bone Mineral Density in Childhood Study is an ongoing longitudinal study in which boys and girls aged 6-16 year old were recruited between 2002-2003, and whose DXA measurements are obtained annually at five clinical centers in the United States ^5; 6^.

BPROOF:

B-PROOF is a trial investigating the effect of 2-year supplementation with 400 mcg folic acid and 500 mcg vitamin B12 on fracture incidence in hyperhomoycsteinemic persons aged 65y and older. The study samples have been previously described in detail [ref: van Wijngaarden JP, Dhonukshe-Rutten RA, van Schoor NM, van der Velde N, Swart KM, Enneman AW, van Dijk SC, Brouwer-Brolsma EM, Zillikens MC, van Meurs JB, Brug J, Uitterlinden AG, Lips P, de Groot LC. Rationale and design of the B-PROOF study, a randomized controlled trial on the effect of supplemental intake of vitamin B12 and folic acid on fracture incidence^7^. All participants gave informed consent and he WU Medical Ethics Committee approved the study protocol, and the Medical Ethics committees of EMC and VUmc gave approval for local feasibility.

Copenhagen Prospective Studies on Asthma (COPSAC) cohort:

The Copenhagen Prospective Studies on Asthma in Childhood is a clinical study. COPSAC_2000_ is a mother child cohort where all the mothers had a history of a doctor’s diagnosis of asthma after 7 years of age and thus this is a high-risk asthma cohort comprising 411 children. Newborns were enrolled in the first month of life, as previously described in detail^8^. COPSAC_2010_ is a mother child cohort comprising 700 children born to unselected mothers from Denmark as described previously in detail^9,10^. The studies were conducted in accordance with the guiding principles of the Declaration of Helsinki and was approved by the Local Ethics Committee (COPSAC2000: KF 01-289/96, COPSAC2010: H-B-2008-093), and the Danish Data Protection Agency (COPSAC2000 and COPSAC2010: 2015-41-3696). Both parents provided written informed consent before enrolment. The Ethics Committee for Copenhagen and the Danish Data Protection Agency approved this study.

deCODE genetics BMD study:

The deCODE genetics BMD study is an ongoing population-based study of all subjects who have undergone a DEXA-Hologic bone mineral density scan at the Landspitali University Hospital, Reykjavik, Iceland. The study samples have been previously described in detail ^11^. All participants gave informed consent and the study was approved by the Data Protection Commission of Iceland and the National Bioethics Committee of Iceland.

ERF:

Erasmus Rucphen Family study (ERF) is a family-based cohort study that includes inhabitants of a genetically isolated community in the South-West of the Netherlands, studied as part of the Genetic Research in Isolated Population (GRIP) program. ERF includes over 3,000 individuals who are living descendants of 22 couples, who had at least six children baptized in the community church, and their spouses. All data were collected between June 2002 and February 2005. The population shows minimal immigration and high inbreeding, therefore frequency of rare alleles is increased in this population. All participants gave informed consent, and the Medical Ethics Committee of the Erasmus University Medical Centre, approved the study.

The Generation R Study:

The Generation R Study is a multiethnic prospective cohort study in which 9,778 pregnant women living in Rotterdam and with delivery date from April 2002 until January 2006 were enrolled. Details of study design and data collection can be found elsewhere ^12^. Genotype and imputation of this cohort are described elsewhere ^13^. The general design, all research aims and the specific measurements in the Generation R Study have been approved by the Medical Ethical Committee of Erasmus MC, University Medical Center Rotterdam. From the age of 12 years onwards, children must sign their own consent form, in accordance with Dutch Law.

GOOD Study:

The Gothenburg Osteoporosis and Obesity Determinants (GOOD) study was initiated to determine both environmental and genetic factors involved in the regulation of bone and fat mass. The GOOD study is a population-based cohort in which male subjects from between 18 and 20 years of age in the Gothenburg area in Sweden were randomly selected using national population registers and invited to participate in this initiative by phone. From the selected candidates 1,068 agreed to participate providing oral and written informed consent. The GOOD study was approved by the local ethics committee at Gothenburg University ^14^.

MROS USA:

The Osteoporotic Fractures in Men (MrOS) Study is a multi-center prospective, longitudinal, observational study of risk factors for vertebral and all non-vertebral fractures in older men, and of the sequelae of fractures in men ^15; 16^. The original specific aims of the study include: (1) to define the skeletal determinants of fracture risk in older men, (2) to define lifestyle and medical factors related to fracture risk, (3) to establish the contribution of fall frequency to fracture risk in older men, (4) to determine to what extent androgen and estrogen concentrations influence fracture risk, (5) to examine the effects of fractures on quality of life, (6) to identify sex differences in the predictors and outcomes of fracture, (7) to collect and store serum, urine and DNA for future analyses as directed by emerging evidence in the fields of aging and skeletal health, and (8) define the extent to which bone mass/fracture risk and prostate diseases are linked. The MrOS Study enrolled 5,994 community dwelling, ambulatory men aged 65 years or older from six communities in the United States (Birmingham, AL; Minneapolis, MN; Palo Alto, CA; Monongahela Valley near Pittsburgh, PA; Portland, OR; and San Diego, CA) between 2000 and 2002. Inclusion criteria were designed to provide a study cohort that is representative of the broad population of older men. The inclusion criteria were: (1) ability to walk without the assistance of another, (2) absence of bilateral hip replacements, (3) ability to provide self-reported data, (4) residence near a clinical site for the duration of the study, (5) absence of a medical condition that (in the judgment of the investigator) would result in imminent death, and (6) ability to understand and sign an informed consent. To qualify as an enrollee, the participant had to provide written informed consent, complete the self-administered questionnaire (SAQ), attend the clinic visit, and complete at least the anthropometric, DXA, and vertebral X-ray procedures. There were no other exclusion criteria. Written informed consent was obtained from all participants, and the Institutional Review Board at each study site approved the study.

Whole body total BMD (g/cm^2^) and head BMD (g/cm^2^) was measured using dual energy x-ray absorptiometry (DXA) (Hologic, Inc., MA) using Hologic QDR 4500 workstations at the baseline clinic visit. A central quality control lab, certification of DXA operators, and standardized procedures for scanning were used to insure reproducibility of DXA measurements. At baseline, a Hologic whole body phantom was circulated and measured at the 6 clinical sites. The variability across clinics was within acceptable limits, and cross-calibration correction factors were not required.

NEO:

The NEO was designed for extensive phenotyping to investigate pathways that lead to obesity-related diseases. The NEO study is a population-based, prospective cohort study that includes 6,671 individuals aged 45–65 years, with an oversampling of individuals with overweight or obesity. At baseline, information on demography, lifestyle, and medical history have been collected by questionnaires. In addition, samples of 24-h urine, fasting and postprandial blood plasma and serum, and DNA were collected. All participants gave written informed consent and the Medical Ethical Committee of the Leiden University Medical Center (LUMC) approved the study.

OPRA:

The Osteoporosis Risk Assessment Cohort (OPRA) cohort recruited Swedish women aged 75, at which time age-related bone loss is already obvious and fractures prevalent. The study was designed to investigate genetic and lifestyle factors contributing to osteoporosis and fracture risk. Of 1604 women invited between December 1995 and May 1999, 1044 (65%) attended at baseline. No exclusion criteria were applied. All participants answered a detailed questionnaire regarding their general health; BMD and body composition was assessed by DXA. All participants gave written informed consent and the Lund University Ethics Committee approved the study.

ORCADES:

The Orkney Complex Disease Study is an ongoing family‐based genetic epidemiology collection in the isolated Scottish archipelago of Orkney. Genetic diversity in this population is decreased compared to Mainland Scotland, consistent with the high levels of endogamy historically. Fasting blood samples were collected and over 300 health-related phenotypes and environmental exposures were measured in each individual. All participants gave informed consent and the study was approved by Research Ethics Committees in Orkney and Aberdeen.

PANIC:

The Physical Activity and Nutrition in Children (PANIC) study is a controlled physical activity and dietary intervention study in a population sample of 506 Finnish children aged 6-8 years at baseline in 2007-2009. Ethical approval was obtained from the Research Ethics Committee of the Hospital District of Northern Savo. All children and their parents gave their written informed consent ^17^. (http://www.uef.fi/en/web/physical-activity-and-nutrition-in-children/home)

RAINE:

The Raine (West Australian Pregnancy Cohort) Study is a longitudinal population-based pregnancy cohort study, which recruited 2,900 pregnant women from the public antenatal clinic at King Edward Memorial Hospital and surrounding private clinics in Perth, Western Australia between May 1989 and November 1991 ^18^. Of the 2868 live births, 1183 had a whole body DXA at 20 years ^19^. The University of Western Australia (UWA) has provided ethical approval.

Rotterdam Study:

The Rotterdam Study is a prospective cohort study of chronic disabling conditions in Dutch elderly individuals that started in 1990 in Ommoord, a suburb of Rotterdam, among 10,994, men and women aged 55 and over ^20^. The Rotterdam Study has been approved by the Medical Ethics Committee of the Erasmus MC (registration number MEC 02.1015) and by the Dutch Ministry of Health, Welfare and Sport (Population Screening Act WBO, license number 1071272-159521-PG). The Rotterdam Study Personal Registration Data collection is filed with the Erasmus MC Data Protection Officer under registration number EMC1712001.

SOF:

The Study of Osteoporotic Fractures (SOF) is a prospective multicenter study of risk factors for vertebral and non-vertebral fractures ^21^. The cohort is comprised of 9,704 community-dwelling women 65 years old or older recruited from populations-based listings in four U.S. areas: Baltimore, Maryland; Minneapolis, Minnesota; Portland, Oregon; and the Monongahela Valley, Pennsylvania. The SOF participants were followed up every four months by postcard or telephone to ascertain the occurrence of falls, fractures and changes in address. To date, follow-up rates have exceeded 95% for vital status and fractures. All fractures are validated by x-ray reports or, in the case of most hip fractures, a review of pre-operative radiographs. The inclusion criteria were: 1) 65 years or older, (2) ability to walk without the assistance of another, (3) absence of bilateral hip replacements, (4) ability to provide self-reported data, (5) residence near a clinical site for the duration of the study, (6) absence of a medical condition that (in the judgment of the investigator) would result in imminent death, and (7) ability to understand and sign an informed consent.

This study used whole body total BMD (g/cm^2^) and head BMD (g/cm^2^) measured using dual energy x-ray absorptiometry (DXA) (Hologic, Inc., MA) using Hologic QDR 2000 workstations at the sixth clinic visit. Scans were performed and analyzed at each clinic. Review of scans was done at the UCSF Coordinating Center on random subsets of scans and on problematic scans identified by technicians at the clinic. Some scans were deemed unacceptable and are not included in the data or are set to a special missing value code.

TwinsUK:

The UK Adult Twin Registry (TwinsUK) (www.twinsuk.ac.uk/) was started in 1993 and is comprised of ~12,000 monozygotic and dizygotic twins (83% female) aged 16-85 years recruited by successive media campaigns from all over the UK without selection for any particular disease or trait. The cohort is from Northern European/UK ancestry and has been shown to be representative of singleton populations and the UK population in general (26). All twins received a series of detailed disease and environmental questionnaires and the majority have been assessed in detail clinically at several time points for several hundred phenotypes related to common diseases or intermediate traits. The primary focus of the study has been the genetic basis of healthy aging process and complex diseases, including cardiovascular, metabolic, musculoskeletal, and ophthalmologic disorders. All participants granted informed consent and favorable ethical opinion was granted by the formerly known St. Thomas’ Hospital Research Ethics Committee (REC).

UKBB:

In 2006-2010, the UK Biobank recruited 502,647 individuals aged between 37-76 years (99.5% were 40-69 years) from across the country. Each participant provided information regarding their health and lifestyle using touch screen questionnaires, physical measurements and agreement to have their health followed and they also provided blood, urine and saliva samples for future analysis. UK Biobank has ethical approval from the Northwest Multi-centre Research Ethics Committee (MREC) and informed consent was obtained from all participants.

### SUPPLEMENTAL ACKNOWLEDGMENTS

The authors would like to thank the many colleagues who contributed to collection and phenotypic characterization of the clinical samples, as well as genotyping and analysis of the GWAS data. Full details of acknowledgements are provided below.

ALSPAC Study:

We are extremely grateful to all the families who took part in this study, the midwives for their help in recruiting them, and the whole ALSPAC team, which includes interviewers, computer and laboratory technicians, clerical workers, research scientists, volunteers, managers, receptionists, and nurses. GWAS data was generated by Sample Logistics and Genotyping Facilities at the Wellcome Trust Sanger Institute and LabCorp (Laboratory Corporation of America) using support from 23andMe. The UK Medical Research Council and the Wellcome Trust (Grant ref: 217065/Z/19/Z) and the University of Bristol provide core support for ALSPAC. This publication is the work of the authors, and JPK and DME will serve as guarantors for the contents of this paper. This work is supported by a Medical Research Council program grant (MC_UU_12013/4 to D.M.E). D.M.E is supported by an NHMRC Senior Research Fellowship (APP1137714). J.P.K is funded by a National Health and Medical Research Council (Australia) Investigator grant (GNT1177938)

The Bone Mineral Density in Childhood Study:

BMDCS is extremely grateful to all the families who participated in this study, and the whole team, which includes interviewers, computer and laboratory technicians, clerical workers, research scientists, volunteers, managers, receptionists, and nurses. This work was funded by the National Institute of Child Health and Human Development (NICHD) contracts NO1-HD-1-3228, -3329, -3330, -3331, -3332 and - 3333, R01 HD058886 and the Clinical and Translational Research Center (5-MO1-RR-000240 and UL1 RR-026314).

BPROOF:

The authors gratefully thank all study participants, and all co-workers who helped to succeed this trial, especially P.H. in ‘t Veld, M. Hillen-Tijdink, A. Nicolaas-Merkus, N. Pliester, S. Oliai Araghi, and S. Smits. They also thank Prof. Dr. H.A.P. Pols for obtaining funding. B-PROOF is supported and funded by The Netherlands Organization for Health Research and Development (ZonMw, Grant 6130.0031), The Hague; unrestricted grant from NZO (Dutch Dairy Association), Zoetermeer; NCHA (Netherlands Consortium Healthy Ageing) Leiden/ Rotterdam; Ministry of Economic Affairs, Agriculture and Innovation (project KB-15-004-003), the Hague; Wageningen University, Wageningen; VU University Medical Center, Amsterdam; Erasmus MC, Rotterdam.

COPSAC:

We express our gratitude to the participants of the COPSAC2000, COPSAC2010 and COPSAC-REGISTRY cohort study for all their support and commitment. We also acknowledge and appreciate the unique efforts of the COPSAC research team.

deCODE:

We thank all the study participants their participation, the staff at deCODE genetics core facilities and the staff at the Research Service Center. deCODE genetics funded the study.

ERF:

We are grateful to all study participants and their relatives, general practitioners and neurologists for their contributions and to P. Veraart for her help in genealogy, J. Vergeer for the supervision of the laboratory work and P. Snijders for his help in data collection. Erasmus Rucphen family study: The ERF study as a part of EUROSPAN (European Special Populations Research Network) was supported by European Commission FP6 STRP grant number 018947 (LSHG-CT-2006-01947) and also received funding from the European Community's Seventh Framework Programme (FP7/2007-2013)/grant agreement HEALTH-F4-2007-201413 by the European Commission under the programme "Quality of Life and Management of the Living Resources" of 5th Framework Programme (no. QLG2-CT-2002-01254). High-throughput analysis of the ERF data was supported by a joint grant from the Netherlands Organization for Scientific Research and the Russian Foundation for Basic Research (NWO-RFBR 047.017.043).

The Generation R Study:

We gratefully acknowledge the contribution of children and parents, general practitioners, hospitals, midwives and pharmacies in Rotterdam. The generation and management of GWAS genotype data for the Generation R Study was done at the Genetic Laboratory of the Department of Internal Medicine, Erasmus MC, The Netherlands. We thank Pascal Arp, Mila Jhamai, Marijn Verkerk, Lizbeth Herrera and Marjolein Peters for their help in creating, managing and QC of the GWAS database. The musculoskeletal research of the Generation R Study is partly supported by the European Commission grant HEALTH-F2-2008-201865-GEFOS. The general design of Generation R Study is made possible by ﬁnancial support from the Erasmus Medical Center, Rotterdam, the Erasmus University Rotterdam, the Netherlands Organization for Health Research and Development (ZonMw), the Netherlands Organisation for Scientific Research (NWO), the Ministry of Health, Welfare and Sport and the Ministry of Youth and Families. Additionally, the Netherlands Organization for Health Research and Development supported authors of this manuscript (ZonMw 907.00303, ZonMw 916.10159 and ZonMw VIDI 016.136.367 to F.R.). J.F.F. has received funding from the European Union’s Horizon 2020 research and innovation programme under grant agreement No 633595 (DynaHEALTH). The study was also supported by funding from the European Union’s Horizon 2020 research and innovation programme (733206, LIFECYCLE).

GOOD Study:

Financial support was received from the Swedish Research Council (K2010- 54X-09894-19-3, 2006-3832 and K2010-52X-20229-05-3),the Swedish Foundation for Strategic Research, the ALF/LUA research grant in Gothenburg, the Lundberg Foundation, the Torsten and Ragnar Söderberg's Foundation, the Västra Götaland Foundation, the Göteborg Medical Society, the Novo Nordisk foundation, and the European Commission grant HEALTH-F2-2008-201865-GEFOS. Financial support was received from the Swedish Research Council (K2010-54X-09894-19-3, 2006-3832 and K2010-52X-20229-05-3), the Swedish Foundation for Strategic Research, the ALF/LUA research grant in Gothenburg, the Lundberg Foundation, the Torsten and Ragnar Söderberg's Foundation, the Västra Götaland Foundation, the Göteborg Medical Society, the Novo Nordisk foundation, and the European Commission grant HEALTH-F2-2008-201865-GEFOS.

MROS-USA:

The Osteoporotic Fractures in Men (MrOS) Study is supported by National Institutes of Health funding. The following institutes provide support: The National Institute on Aging (NIA), the National Institute of Arthritis and Musculoskeletal and Skin Diseases (NIAMS), the National Center for Advancing Translational Sciences (NCATS), and NIH Roadmap for Medical Research under the following grant numbers: U01 AG027810, U01 AG042124, U01 AG042139, U01 AG042140, U01 AG042143, U01 AG042145, U01 AG042168, U01 AR066160, and UL1 TR000128. NIAMS provided funding for the MrOS ancillary study ‘Replication of candidate gene associations and bone strength phenotype in MrOS’ under the grant number R01 AR051124 and the MrOS ancillary study ‘GWAS in MrOS and SOF’ under the grant number RC2 AR058973.

NEO:

The authors of the NEO study thank all individuals who participated in the Netherlands Epidemiology in Obesity study, all participating general practitioners for inviting eligible participants and all research nurses for collection of the data. We thank the NEO study group, Pat van Beelen, Petra Noordijk and Ingeborg de Jonge for the coordination, lab and data management of the NEO study. The genotyping in the NEO study was supported by the Centre National de Génotypage (Paris, France), headed by Jean-Francois Deleuze. The NEO study is supported by the participating Departments, the Division and the Board of Directors of the Leiden University Medical Center, and by the Leiden University, Research Profile Area Vascular and Regenerative Medicine. Dennis Mook-Kanamori is supported by Dutch Science Organization (ZonMW-VENI Grant 916.14.023).

OPRA:

This work was supported by grants from the Swedish Research Council (K2015-52X-14691-13-4), Forte (Grant 2007–2125), Greta and Johan Kock Foundation, A. Påhlsson Foundation, A. Osterlund Foundation, H Järnhardt foundation, King Gustav V 80 year fund, King Gustav V and Queen Victoria Foundation, Åke Wiberg Foundation, Thelma Zoegas Foundation, Swedish Rheumatism foundation, Skåne University Hospital Research Fund and the Research and Development Council of Region Skåne, Sweden.

Thanks are extended to the research nurses at the Clinical and Molecular Osteoporosis Research Unit, Malmö; Åsa Almgren for data management, Olle Melander for GWAS genotyping (performed at Lund University, Dept of Clinical Sciences Malmö, Sweden) and to all the women who kindly participated in the study.

ORCADES:

ORCADES was supported by the Chief Scientist Office of the Scottish Government (CZB/4/276, CZB/4/710), the Royal Society, the MRC Human Genetics Unit, Arthritis Research UK and the European Union framework program 6 EUROSPAN project (contract no. LSHG-CT-2006-018947). DNA extractions were performed at the Wellcome Trust Clinical Research Facility in Edinburgh. We would like to acknowledge the invaluable contributions of the research nurses in Orkney, the administrative team in Edinburgh and the people of Orkney.

PANIC:

We are grateful to the parents and children for participating in the PANIC study and to the whole research team for their contribution in carrying out the study. The PANIC study was financially supported by grants from Ministry of Social Affairs and Health of Finland, Ministry of Education and Culture of Finland, Finnish Innovation Fund Sitra, Social Insurance Institution of Finland, Finnish Cultural Foundation, Juha Vainio Foundation, Foundation for Pediatric Research, Paavo Nurmi Foundation, Paulo Foundation, Diabetes Research Foundation, Yrjo Jahnsson Foundation, Finnish Foundation for Cardiovascular Research, State Research Funding from Research Committee of Kuopio University Hospital Catchment Area, Kuopio University Hospital EVO Funding, National Doctoral Programs, and the City of Kuopio.

RAINE:

We thank the Raine Study participants and families for being part of the study, the Raine Study Team for cohort management and data collection. The Raine Study receives core funding support from the University of Western Australia -Faculty of Medicine, Dentistry and Health Sciences, Curtin University, the Raine Medical Research Foundation, the Women and Infants Research Foundation, Telethon Kids Institute and Edith Cowan University. The study was funded by the National Health and Medical Research Council of Australia [grant numbers 572613, 403981 and 003209], the Canadian Institutes of Health Research [grant number MOP-82893] and the Lions Eye Institute in WA. This work was supported by resources provided by the Pawsey Supercomputing Centre with funding from the Australian Government and the Government of Western Australia.

Rotterdam Study:

The generation and management of GWAS genotype data for the Rotterdam Study (RS I, RS II, RS III) was executed by the Human Genotyping Facility of the Genetic Laboratory of the Department of Internal Medicine, Erasmus MC, Rotterdam, The Netherlands. The GWAS datasets are supported by the Netherlands Organisation of Scientific Research NWO Investments (nr. 175.010.2005.011, 911-03-012), the Genetic Laboratory of the Department of Internal Medicine, Erasmus MC, the Research Institute for Diseases in the Elderly (014-93-015; RIDE2), the Netherlands Genomics Initiative (NGI)/Netherlands Organisation for Scientific Research (NWO) Netherlands Consortium for Healthy Aging (NCHA), project nr. 050-060-810. We thank Pascal Arp, Mila Jhamai, Marijn Verkerk, Lizbeth Herrera and Marjolein Peters, MSc, and Carolina Medina-Gomez, MSc, for their help in creating the GWAS database, and Karol Estrada, PhD, Yurii Aulchenko, PhD, and Carolina Medina-Gomez, PhD, for the creation and analysis of imputed data. We would like to thank Dr. Karol Estrada, Dr. Fernando Rivadeneira, Dr. Tobias A. Knoch, Marijn Verkerk, Anis Abuseiris, Dr. Linda Boer and Rob de Graaf (Erasmus MC Rotterdam, The Netherlands), for their help in creating and maintaining GRIMP. Dr. Fernando Rivadeneira received an additional grant from the Netherlands Organization for Health Research and Development ZonMw VIDI 016.136.367. The Rotterdam Study is funded by Erasmus Medical Center and Erasmus University, Rotterdam, Netherlands Organization for the Health Research and Development (ZonMw), the Research Institute for Diseases in the Elderly (RIDE), the Ministry of Education, Culture and Science, the Ministry for Health, Welfare and Sports, the European Commission (DG XII), and the Municipality of Rotterdam. The authors are very grateful to the study participants, the staff from the Rotterdam Study (particularly L. Buist and J.H. van den Boogert) and the participating general practitioners and pharmacists.

SOF:

The Study of Osteoporotic Fractures (SOF) is supported by National Institutes of Health funding. The National Institute on Aging (NIA) provides support under the following grant numbers: R01 AG005407, R01 AR35582, R01 AR35583, R01 AR35584, R01 AG005394, R01 AG027574, and R01 AG027576. The National Institute of Arthritis and Musculoskeletal and Skin Diseases (NIAMS) provides funding for the SOF ancillary study ‘GWAS in MrOS and SOF’ under the grant number RC2AR058973.

Twins UK:

We gratefully acknowledge the help and support of the twin volunteers. TwinsUK is funded by the Wellcome Trust, Medical Research Council, European Union, the National Institute for Health Research (NIHR)-funded BioResource, Clinical Research Facility and Biomedical Research Centre based at Guy’s and St Thomas’ NHS Foundation Trust in partnership with King’s College London. TDS is holder of an ERC Advanced Principal Investigator award. SNP Genotyping was performed by The Wellcome Trust Sanger Institute and National Eye Institute via NIH/CIDR for TwinsUK. The study also received support from the Australian National Health and Medical Research Council (Project Grant 1048216) and The Pawsey Supercomputing Centre (with Funding from the Australian Government and the Government of Western Australia; PG 16/0162, PG 17/director2025).

Functional Group:

C Ackert-Bicknell: National Institute of Health /National Institute of Arthritis Musculoskeletal and Skin Diseases grant number AR060981.
